# Triglyceride-glucose index as a biomarker to differentiate stroke subtypes: A hospital-based cross-sectional study

**DOI:** 10.1101/2023.08.30.23294872

**Authors:** Nizar Daoussi, Imen Zemni, Yasmin Saad, Amal Abbes, Rihab Ben Dhia, Mariem Mhiri, Asma Belghith-Sriha, Mahbouba Frih-Ayed

**Affiliations:** Neurology Department, University Hospital of Monastir, Av 1^st^ June 5000 Monastir; Community Health and Epidemiology Department, University Hospital of Monastir, Av 1^st^ June 5000 Monastir

## Abstract

**Background:** A growing body of literature suggests that the triglyceride-glucose (TyG) index is linked to ischemic stroke in several ways. The usefulness of this biomarker to differentiate etiologic stroke subtypes has not been thoroughly studied. We aimed to figure out whether the TyG index differentiates cardioembolic (CE) from non-cardioembolic (NCE) strokes.

**Methods:** A cross-sectional hospital-based study of consecutive stroke cases admitted to the University Hospital of Monastir in Tunisia from January 2018 to December 2022. The TyG index was calculated through the natural logarithm (Ln) of the product of triglyceride and glucose fasting levels. A binary logistic regression was performed to analyze the association between the TyG index and the studied stroke subtypes. We plotted the receiver operating characteristic curve (ROC curve) to determine the best cutoff point for the TyG index in differentiating between CE and NCE stroke subtypes.

**Results:** We included 320 patients (mean age: 64.2 ±11.1 years; 65.3% males). The TyG index values were independently associated with the NCE subtype (OR=2.38; 95% CI=1.52-3.73; p<0.001) when analyzed as a continuous data variable. Logistic regression of quartile distribution showed that the probability of developing a NCE stroke increased proportionally with the TyG index quartiles. The ROC curve showed an area under the curve of 0.636 (95% CI=0.565-0.707; p<0.001) with a cutoff of 8.8 (sensitivity = 68.8%, specificity = 57%).

**Conclusion:** High levels of the TyG index are associated with a higher prevalence of NCE stroke while low values are associated with CE strokes. Thus, the TyG index can be a useful biomarker in the differentiation between CE and NCE stroke subtypes.

## Introduction

Stroke is a leading cause of disability and death worldwide. This condition can be preventable provided the cause is recognized. Ischemic stroke etiology is generally established to initiate appropriate secondary prevention and avoid the recurrence of events. However, this can be challenging during the acute phase of stroke due to limited access to investigations in many healthcare facilities, especially in developing countries. Insulin resistance (IR) is proven to be a major vascular risk factor regardless of the patient’s diabetes status and independently from other risk factors^1^. The triglyceride-glucose (TyG) index is a simple, reliable, and inexpensive surrogate marker of IR calculated from fasting levels of triglycerides and glucose^2^. A growing body of literature suggests that the TyG index is linked to ischemic stroke in several ways. A recent meta-analysis showed that a higher TyG index level was independently associated with a greater risk of first-ever stroke^3^. Moreover, increasing the TyG index was associated with a high risk of early neurologic deterioration after stroke^4^. Several other studies showed an elevated risk of stroke recurrence^5^ and mortality^6^ in patients with a high TyG index. The usefulness of this biomarker to differentiate etiologic stroke subtypes has not been thoroughly studied. The purpose of our study was to figure out whether the TyG index differentiates cardioembolic (CE) from non-cardioembolic (NCE) strokes.

## Methods

### Study design and participants

We conducted a cross-sectional hospital-based study of consecutive stroke cases admitted to the University Hospital of Monastir in Tunisia from January 2018 to December 2022. Patients who arrived at the emergency department with suspected stroke and underwent CT scans were subsequently admitted to the hospital and included in the study. The diagnosis of stroke was reviewed and confirmed by a senior trained neurologist. Individuals diagnosed with hemorrhagic stroke or subarachnoid hemorrhage were not included in this study. We also excluded patients with transient ischemic attacks, embolic stroke of undetermined source, strokes of other determined causes, and those with missing data or blood tests collected beyond 24 hours from stroke onset. Data were collected from numeric medical records including relevant history and vascular risk factors. The clinical severity of stroke was assessed by using the National Institutes of Health Stroke Scale (NIHSS score). A routine laboratory investigation including fasting glucose, lipid profile, and glycosylated hemoglobin was collected on the first day of admission. All included individuals had an etiologic workup including carotid Doppler or CT angiography for carotid and intracranial atheroma, electrocardiogram(ECG), 24-hour Holter ECG, and transthoracic echography for CE sources. Stroke subtypes were categorized according to the Trial of ORG 10172 in Acute Stroke Treatment (TOAST) classification. We analyzed two groups of patients: those with confirmed CE sources and those with NCE origin, including small vessel disease (SVD) and large artery atheroma (LAA). The ethics committee of the University Hospital of Monastir approved the study. The STROBE (Strengthening the Reporting of Observational Studies in Epidemiology) reporting guidelines (https://www.strobe-statement.org/) were followed in this study report.

### TyG index assessment

We determined the TyG index through the natural logarithm (Ln) of the product of triglyceride and glucose levels (2). Fasting blood samples were obtained from all patients during the first 24 hours of their hospital admission. Patients included in the study were divided into four groups ranging from quartile 1 to quartile 4 according to their TyG index values.

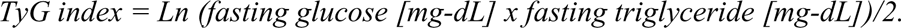

### Data analysis

Qualitative variables were expressed as numbers and percentages while quantitative variables were presented by mean and standard deviation or median and interval interquartile after assessing normality with the Kolmogorov-Smirnov test. Pearson chi2 or Fischer exact tests were used for the comparison of qualitative variables. Continuous data were compared using the T-test for independent samples or the Mann-Whitney U test, as appropriate. We considered the result statistically significant if the p-value was less than 0.05. A binary logistic regression was performed to analyze the association between the TyG index and the studied stroke subtypes. The sample was sorted into four quartiles based on the TyG index values. The ANOVA test was conducted to compare subtypes of stroke among the quartiles of TyG index values. We also plotted the receiver operating characteristic curve (ROC curve) to determine the best cutoff point for the TyG index in differentiating between CE and NCE subtypes according to Youden’s index. The SPSS statistical software package, SPSS version 22.0 (SPSS Inc., Chicago, IL) was used for statistical analysis.

## Results

### Baseline characteristics and comparison between NCE and CE groups

We initially screened 479 patients for eligibility in this study. After applying exclusion criteria, only 320 patients were included. The TOAST classification was used to categorize the sample into different groups based on the etiology as follows: patients with SVD (118 patients), LAA including intra and extracranial stenosis (117 patients), CE origin (85 patients), undetermined (98 patients) and other determined etiologies (5 patients). Patients included in the analysis were divided into two groups: one group with confirmed CE sources while the other consisted of patients with NCE origin, including SVD and LAA (Fig. 1).

**Fig.1.**
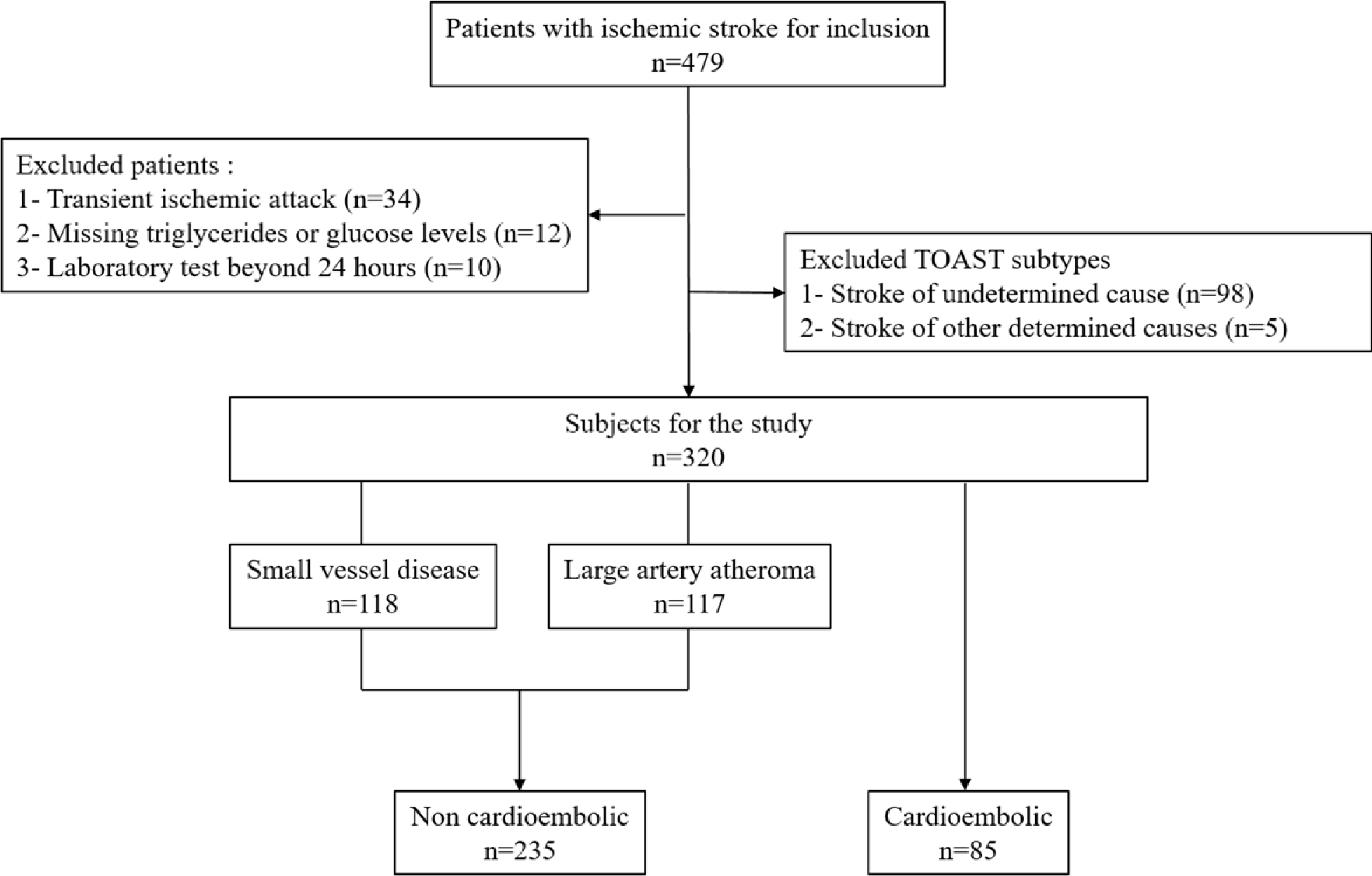
Flowchart of included patients.

The mean age of the study participants was 64.2 years (SD, 11.1) and 209 (65.3%) patients were males (Table 1). Patients diagnosed with NCE stroke were significantly younger (62.4 ±9.7; p<0.001) and had a higher rate of males (73.6%; p<0.001) compared to patients in the CE group. Additionally, the NCE group had a higher prevalence of smokers (41.7%; p=0.001). In contrast, patients in the CE group had a more severe clinical presentation with higher NIHSS scores (8.65 ±5.9; p<0.001) and greater occurrence of consciousness disturbance (16.4%; p<0.001). The lipid profile components were found to be elevated in NCE compared to CE strokes. The TyG index values (9.17 ±0.68; p<0.001) were significantly associated with the NCE subtype.

**Table 1.**
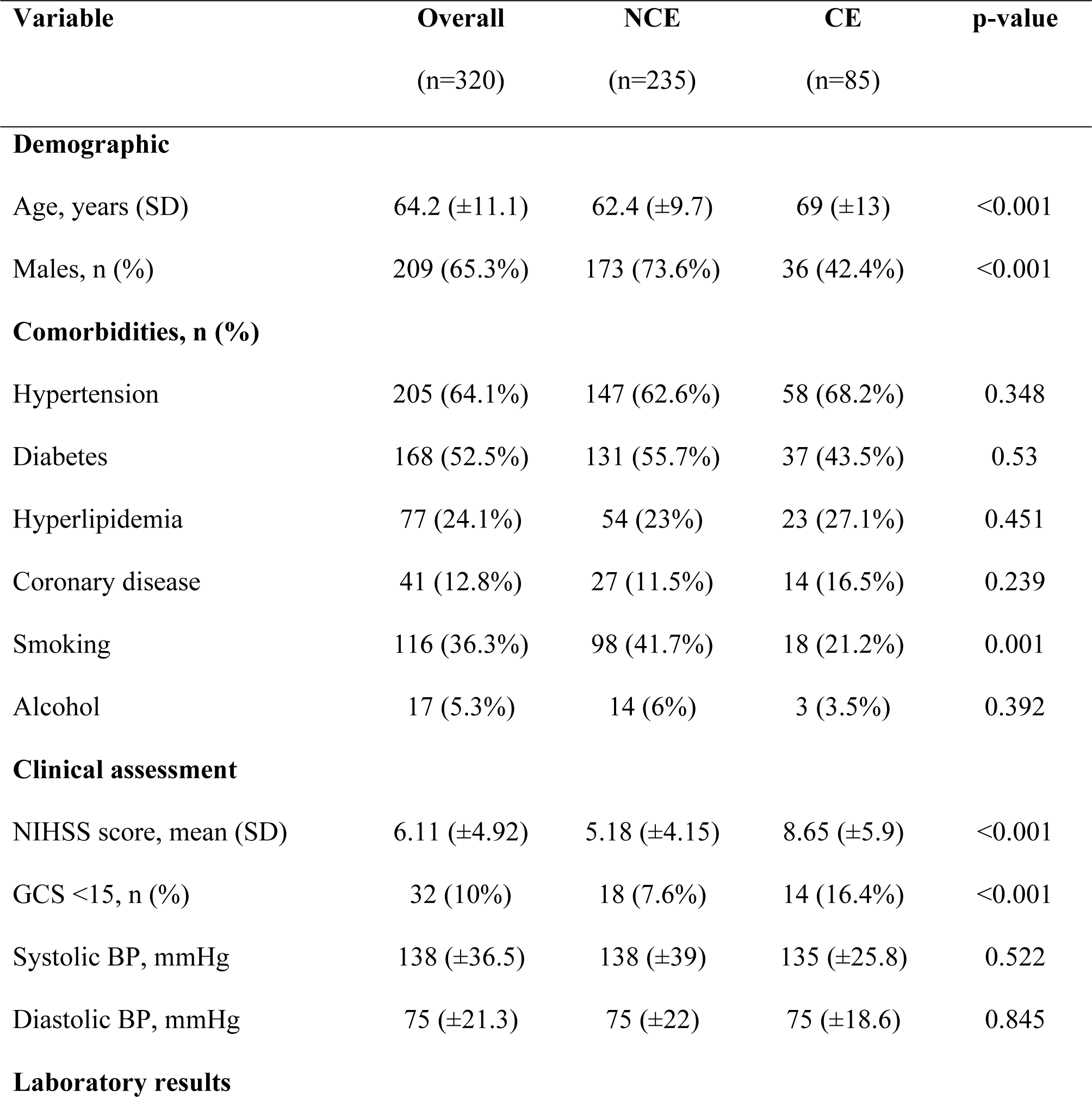
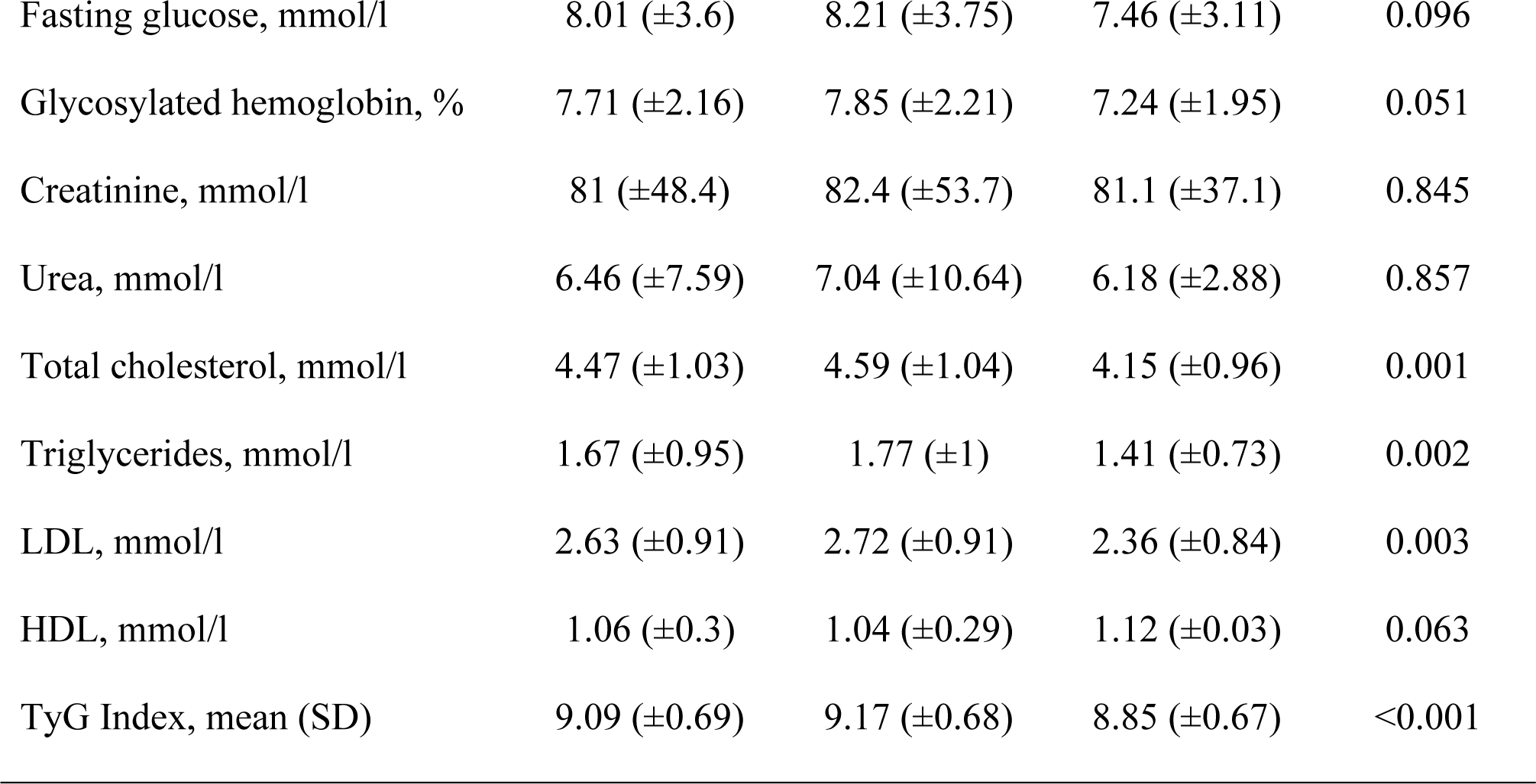
Characteristics according to the stroke studied subtypes.

| Variable | Overall<br>(n=320) | NCE<br>(n=235) | CE<br>(n=85) | p-value |
| --- | --- | --- | --- | --- |
| **Demographic** |  |  |  |  |
| Age, years (SD) | 64.2 ($\pm 11.1$) | 62.4 ($\pm 9.7$) | 69 ($\pm 13$) | <0.001 |
| Males, n (%) | 209 (65.3%) | 173 (73.6%) | 36 (42.4%) | <0.001 |
| **Comorbidities, n (%)** |  |  |  |  |
| Hypertension | 205 (64.1%) | 147 (62.6%) | 58 (68.2%) | 0.348 |
| Diabetes | 168 (52.5%) | 131 (55.7%) | 37 (43.5%) | 0.53 |
| Hyperlipidemia | 77 (24.1%) | 54 (23%) | 23 (27.1%) | 0.451 |
| Coronary disease | 41 (12.8%) | 27 (11.5%) | 14 (16.5%) | 0.239 |
| Smoking | 116 (36.3%) | 98 (41.7%) | 18 (21.2%) | 0.001 |
| Alcohol | 17 (5.3%) | 14 (6%) | 3 (3.5%) | 0.392 |
| **Clinical assessment** |  |  |  |  |
| NIHSS score, mean (SD) | 6.11 ($\pm 4.92$) | 5.18 ($\pm 4.15$) | 8.65 ($\pm 5.9$) | <0.001 |
| GCS <15, n (%) | 32 (10%) | 18 (7.6%) | 14 (16.4%) | <0.001 |
| Systolic BP, mmHg | 138 ($\pm 36.5$) | 138 ($\pm 39$) | 135 ($\pm 25.8$) | 0.522 |
| Diastolic BP, mmHg | 75 ($\pm 21.3$) | 75 ($\pm 22$) | 75 ($\pm 18.6$) | 0.845 |
| **Laboratory results** |  |  |  |  |
| Fasting glucose, mmol/l | 8.01 (±3.6) | 8.21 (±3.75) | 7.46 (±3.11) | 0.096 |
| Glycosylated hemoglobin, % | 7.71 (±2.16) | 7.85 (±2.21) | 7.24 (±1.95) | 0.051 |
| Creatinine, mmol/l | 81 (±48.4) | 82.4 (±53.7) | 81.1 (±37.1) | 0.845 |
| Urea, mmol/l | 6.46 (±7.59) | 7.04 (±10.64) | 6.18 (±2.88) | 0.857 |
| Total cholesterol, mmol/l | 4.47 (±1.03) | 4.59 (±1.04) | 4.15 (±0.96) | 0.001 |
| Triglycerides, mmol/l | 1.67 (±0.95) | 1.77 (±1) | 1.41 (±0.73) | 0.002 |
| LDL, mmol/l | 2.63 (±0.91) | 2.72 (±0.91) | 2.36 (±0.84) | 0.003 |
| HDL, mmol/l | 1.06 (±0.3) | 1.04 (±0.29) | 1.12 (±0.03) | 0.063 |
| TyG Index, mean (SD) | 9.09 (±0.69) | 9.17 (±0.68) | 8.85 (±0.67) | <0.001 |

### TyG index and stroke subtypes

A binary logistic regression was used to identify independent factors associated with CE and NCE stroke subtypes. The TyG index values were independently associated with the NCE subtype (OR=2.38; 95% CI=1.52-3.73; p<0.001) when analyzed as a continuous data variable (Table 2). Meanwhile, older age (OR=0.33; 95%CI=0.16-0.64; p=0.001), male gender (OR=0.95; 95% CI=0.93-0.98; p=0.002), and clinical severity of stroke (OR=0.88; 95% CI=0.82-0.94; p<0.001) were found to be independent factors linked to the CE subtype.

**Table 2.**
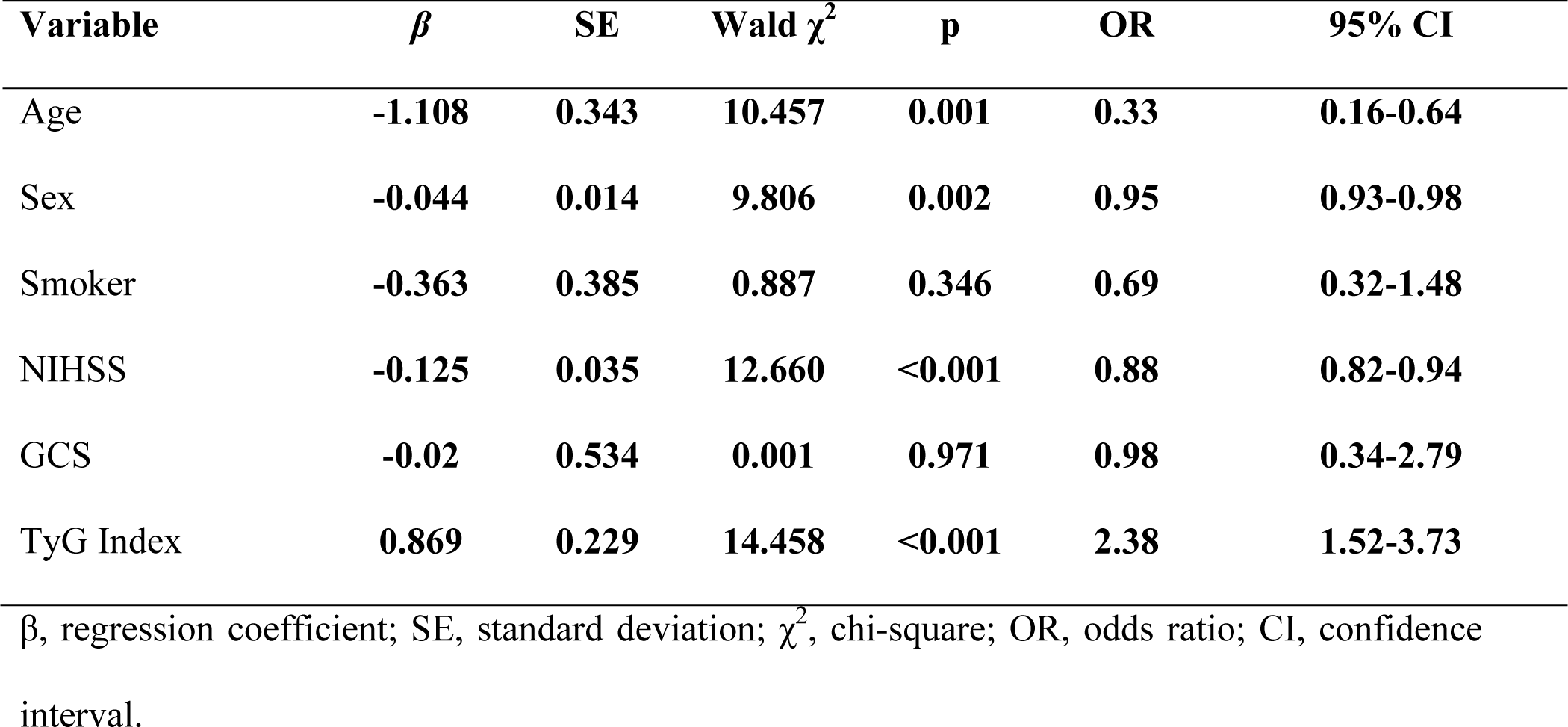
Binary logistic regression for CE versus NCE subtypes: Model 1.

To achieve greater accuracy in studying the link between the TyG index and stroke subtypes, the sample was divided equally into four quartiles (Table 3) of 80 patients based on the TyG index values: Q1 [7.22-8.55], Q2 [8.56-9.11], Q3 [9.12-9.53], and Q4 [9.54-11.62]. As shown in Figure 2, there was a greater likelihood of NCE strokes occurring with a higher TyG index, whereas a lower TyG index was associated with an increased frequency of CE strokes.

**Table 3.** Binary logistic regression for TyG index quartiles: Model 2.

| Variable | $\beta$ | SE | Wald $\chi^2$ | P | OR | 95% CI |
| --- | --- | --- | --- | --- | --- | --- |
| Quartile 1 |  |  | 16.997 | 0.001 |  |  |
| Quartile 2 | 0.982 | 0.394 | 6.222 | 0.013 | 2.66 | 1.23-5.77 |
| Quartile 3 | 1.080 | 0.407 | 7.059 | 0.008 | 2.94 | 1.32-6.53 |
| Quartile 4 | 1.802 | 0.458 | 15.465 | 0.000 | 6.06 | 2.46-14.87 |

$\beta$ , regression coefficient; SE, standard deviation; $\chi^2$ , chi-square; OR, odds ratio; CI, confidence interval. Adjusted for: age, sex, smoking, NIHSS, and GCS scores.

**Fig. 2.**
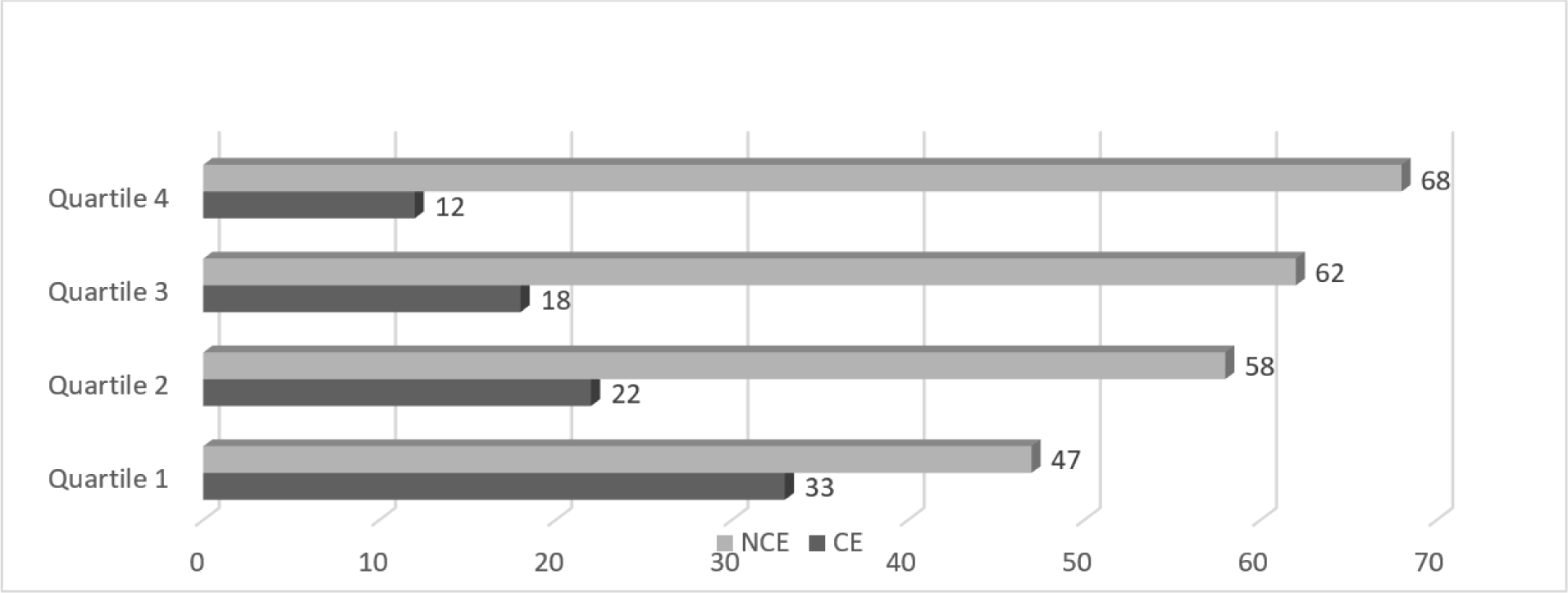
Distribution of CE and NCE subtypes according to the TyG index quartiles.

We performed a second model of logistic regression in which we included the TyG index as a categorical variable based on quartile distribution and we found that the probability of developing a NCE stroke increased proportionally with the TyG index quartiles (Table 3).

### The discriminatory ability of the TyG index in CE versus NCE subtypes

We plotted the ROC curve to calculate the sensitivity and specificity of the TyG index in the differentiation between the CE and NCE stroke subtypes (Fig. 3). The area under the curve (AUC) was 0.636 (95% CI = 0.565-0.707; p<0.001). According to the Youden index, the best TyG index cutoff in differentiating between CE and NCE was 8.8 with a sensitivity of 68.8% and specificity of 57%.

**Fig. 3.**
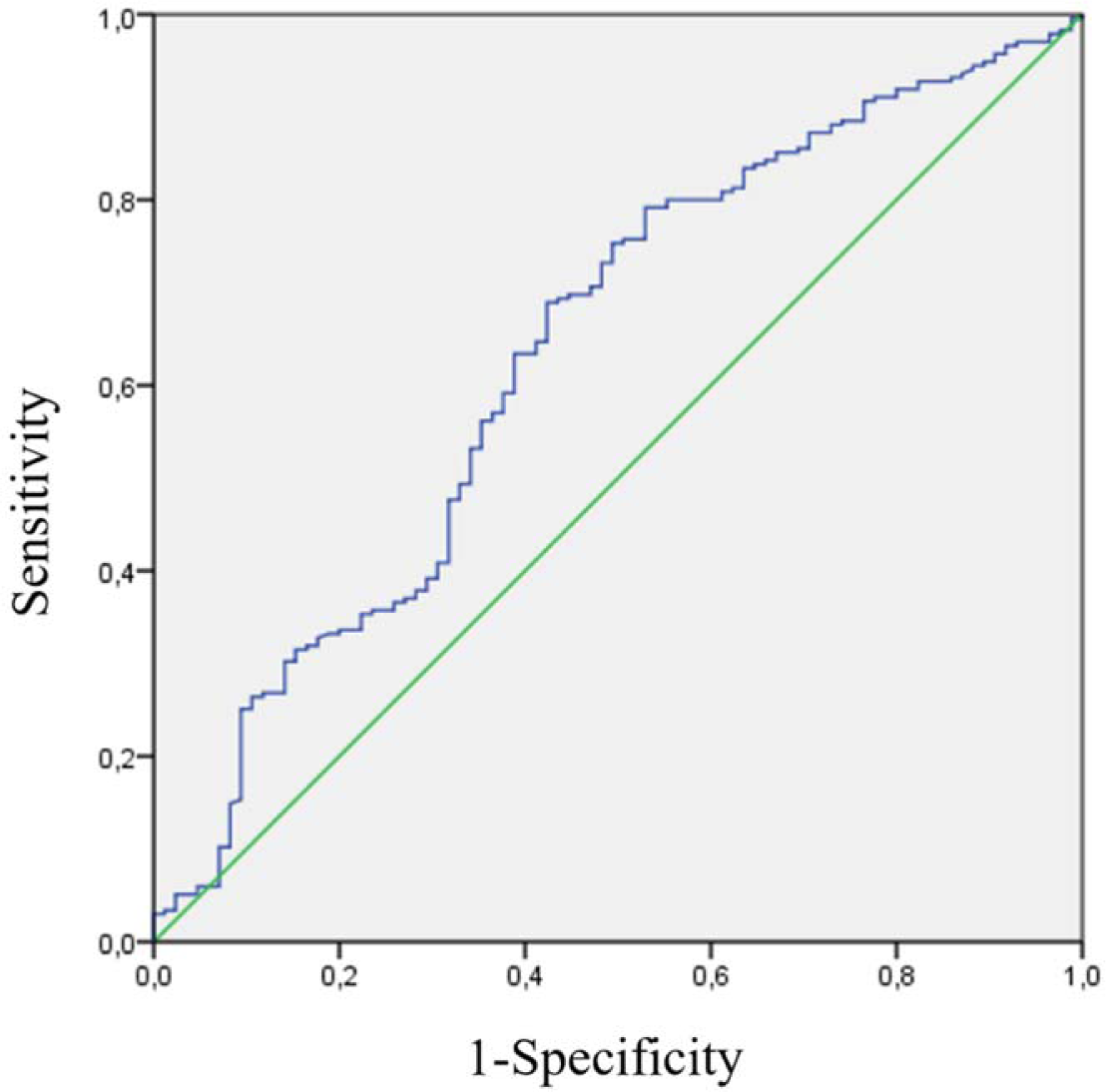
ROC curve of the TyG index for differentiation between CE and NCE subtypes.

## Discussion

Our main finding is that the TyG index was positively correlated with the NCE strokes and inversely related to the CE subtype of stroke. Even after adjusting for other confounders, the association of the TyG index with NCE stroke remained strong (OR=2.38; 95% CI=1.52-3.73). In the absence of a defined cutoff value for the TyG index, we divided the sample into quartiles and analyzed the TyG index as a categorical variable. We observed a gradual increase in the odds ratio of being in the NCE group with higher TyG index values (4th Quartile OR= 6.06; 95% CI=2.46-14.87). These findings suggest a dose-dependent relationship between the TyG index values and CE/NCE stroke subtypes.

IR, represented by the TyG index, is known to be a vascular risk factor for cardiovascular diseases^7^. Chen et al demonstrated that genetically predicted IR phenotypes increase ischemic stroke risk, in particular the SVD and potentially LAA subtype. However, they found no significant association of IR with the CE subtype^8^. The underlying mechanism responsible for ischemic stroke includes several factors: activation of vascular endothelial sodium channels resulting in vascular stiffness and hypertension^9^, systemic inflammation, and atherogenesis^10^. Miao et al demonstrated a linear correlation between the TyG index and the abnormal intima-media thickness and the existence of carotid plaques^11^. Additionally, the increase in the TyG index was reported to be related to the acceleration of carotid plaque instability^12^. Atherosclerosis is the primum-movens of SVD, intracranial atheroma, and LAA. It is responsible for perforating artery occlusion in SVD and artery-to-artery embolism or vessel stenosis and occlusion in both LAA and intracranial atheroma. Thus, we considered these subtypes of stroke in the same group and excluded from our study strokes of undetermined cause and strokes of other etiologies, as the mechanism is not established. Several studies were carried out to understand the link between the TyG index and stroke subtypes. Zhang et al reported that the TyG index is correlated with SVD and may have the potential to optimize the risk stratification of SVD^13, 14^. Furthermore, the TyG index was reported to be a useful biomarker of LAA stroke and can help in identifying candidates for carotid reperfusion^15^. A recent study demonstrated that the TyG index is a significant risk factor for intracranial stenosis and has a linear association with the number and degree of vessel stenosis^16^. On the other hand, none of the previous reports mentioned a high TyG index in the CE stroke subtype.

In our study, the optimal cutoff point of the TyG index to differentiate between CE and NCE stroke subtypes was 8.8. Several studies have investigated the most effective threshold of the TyG index depending on the specific outcome studied. For example, a cutoff of 8.4 was reported to differentiate patients with intracranial stenosis from patients with minor stroke and hypertension^16^. Furthermore, the risk of post-stroke mortality was significantly increased for patients with a TyG index >9.13^6^. In patients with SVD, a cutoff of 9.01 was associated with cognitive impairment in diabetic elderly individuals^17^. A TyG index value higher than 9.06 was reported to be associated with a higher risk of carotid plaque^18^.

To the best of our knowledge, this is the first report investigating the ability of the TyG index to differentiate CE from NCE stroke subtypes. However, our study had several limitations. First, the cross-sectional methodology, cannot exclude the possibility of residual confounders. As a consequence, the causal relationship between the TyG index and the studied stroke subtypes cannot be established. Therefore, further prospective cohort studies are needed to confirm our findings. Second, previous use of antidiabetic and lipid-lowering medications was not considered in our study. This may affect the TyG index values and interfere with the discriminatory ability of the TyG index in stroke subtypes. Third, performing the study in a unique center limits the generalizability of our results.

## Conclusion

High levels of the TyG index are associated with a higher prevalence of NCE stroke while low values are associated with CE strokes. Thus, the TyG index can be a useful biomarker in the differentiation between CE and NCE stroke subtypes. Further multicenter prospective cohort studies are needed to confirm this finding and establish a possible causality link between the TyG index and stroke etiologies.

## Data Availability

The data that support the findings of this study are available on request from the corresponding author, ND

## Nonstandard abbreviations and acronyms

CE: Cardioembolic
IR: Insulin resistance
Ln: Natural logarithm
LAA: Large artery atheroma
NIHSS: National Institutes of Health Stroke Scale
NCE: Non cardioembolic
SVD: Small vessel disease
TOAST: Trial of ORG 10172 in Acute Stroke Treatment
TyG: Triglyceride-Glucose Index

